# Evidence of supratentorial white matter injury prior to treatment in children with posterior fossa tumours using diffusion MRI

**DOI:** 10.1101/2024.12.16.24318543

**Authors:** Emily R. Drabek-Maunder, Jenny Gains, Darren R. Hargrave, Kshitij Mankad, Kristian Aquilina, Jamie A. Dean, Andrew Nisbet, Chris A. Clark

## Abstract

**Background:** Paediatric brain tumour survivors can have neurocognitive deficits that negatively impact their quality of life, but it is unclear if deficits are primarily caused by treatments, such as radiotherapy, or manifest earlier due to the tumour and related complications. The aim of this work is to characterise white matter injury caused by brain tumours, unrelated to treatment effects, and explore heterogeneity in these white matter abnormalities between individual patients.

**Methods:** We used diffusion tensor imaging (DTI) and Neurite Orientation Dispersion Diffusion Imaging (NODDI) to assess white matter injury in 8 posterior fossa tumour patients. A novel one-against-many approach was used by comparing an individual patient to 20 age- and sex-matched healthy controls to assess variability in white matter abnormalities between the posterior fossa tumour patients. White matter was analysed at presentation (prior to treatment), post-surgery (24-72 hours after surgery), and at follow-up (3-18 months after surgery).

**Results:** We demonstrate white matter abnormalities in 5 posterior fossa tumour patients before treatment, likely related to tumour-induced hydrocephalus, which persisted after treatment. White matter changes were complex and patient-specific, and group-based comparisons with control subjects may fail to detect these individual abnormalities.

**Conclusions:** Identifying pre-treatment white matter injury in posterior fossa tumour patients highlights the importance of personalised assessment of brain microstructure, which should be considered in minimising neurocognitive deficits to improve patient quality of life.

## 1 Introduction

Over half of all childhood brain tumours originate in the posterior fossa [1]. These include pilocytic astrocytoma (WHO Grade I), a slow-growing, low-grade tumour; ependymoma (WHO Grade II/III); and medulloblastoma (WHO Grade IV), the most common malignant brain tumour in childhood [2, 3]. Treatment for posterior fossa tumours typically involves surgery, followed by chemotherapy and/or radiotherapy depending on the histology of the tumour and the age of the patient. Survivors face a range of neuropsychological impairments, which impact behaviour, cognition, language and motor skills [4–7]. Neurological outcomes are associated with white matter changes within the brain [8–13], but it is unclear whether injury to white matter occurs as a result of treatment, such as radiotherapy, or is already present at diagnosis, for example secondary to hydrocephalus [14–16].

It is possible that white matter damage observed prior to treatment can contribute to neurocognitive deficits experienced after treatment. To improve treatment strategies and patient outcomes, it is important to understand the cause of these white matter abnormalities and any potential cumulative injury over time. A non-invasive method that can be used to investigate white matter microstructure is diffusion-weighted magnetic resonance imaging (diffusion MRI), which quantifies the motion of water molecules within the brain (i.e. water diffusion).

The diffusion of water molecules can be modelled using different methods. One of the most common techniques is diffusion tensor imaging (DTI), which models the magnitude and directionality of diffusion using the diffusion tensor, a symmetric 3*×*3 matrix of the magnitude of water diffusion in 3 dimensions [17]. Water diffusion is restricted by the underlying tissue microstructure. Therefore, properties of the diffusion tensor can be used to infer information about abnormalities in the structure of white matter, such as myelination, axon organisation, oedema, gliosis, density and dispersion [18–21]. DTI parameters that can be measured include: fractional anisotropy (FA), the measurement of directionality or the degree of anisotropy of the water diffusion; mean diffusivity (MD), the magnitude of water diffusion; axial diffusivity (AD), the magnitude of diffusion parallel to the principle diffusion direction; radial diffusivity (RD), the magnitude of diffusion perpendicular to the principle diffusion direction. The most common parameter used to assess white matter microstructure in DTI studies is FA. Diffusion in white matter is expected to be more anisotropic, aligning along the white matter tracts.

Using DTI, previous studies have investigated the cause of white matter changes in posterior fossa tumour patients by comparing patient groups with different treatment types or radiation doses, analysing the impact of tumour size or location, and exploring longitudinal changes after surgery and adjuvant treatments [8, 13–16, 22–30]. However, DTI has not been used to quantify tumour-associated white matter damage in posterior fossa tumour patients prior to treatment. Variability between patients has also not been previously explored, for example, by assessing white matter in a ‘one-against-many’ approach, which is a technique that compares an individual patient to age- and sex-matched controls (as opposed to comparing a patient group to a matched control group).

While DTI is valuable for modelling water diffusion in the brain, the model is limited by its lack of specificity. For example, changes in the anisotropy (FA) of water diffusion could result from changes in the coherence of white matter tracts and the degree to which axon bundles are aligned, or properties of the tracts themselves (axon density, diameter or myelination). More advanced diffusion models have been developed to address limitations in the DTI model, including the Neurite Orientation Dispersion Diffusion Imaging (NODDI), which enables the measurement of axon density and dispersion [31].

We sought to use DTI and NODDI to assess white matter damage in paediatric patients with posterior fossa tumours prior to any treatment. We focus on comparing individual brain tumour patients to age- and sex-matched controls in a one-against-many approach to identify if patients have white matter damage due to the tumour and its related effects prior to any treatment.

## 2 Methods

### 2.1. Participants

This study included 8 paediatric patients with posterior fossa tumours treated at Great Ormond Street Hospital for Children (GOSH) (Table 1). In all patients, surgery was their first intervention; all patients underwent MR imaging prior to surgery (Table 2). Six patients received further imaging at a time point directly after surgery (‘post-surgery’ or 24-72 hours after tumour resection) and/or at an early follow-up (‘follow-up’ or 3 months after surgical resection). Two patients had further follow-up imaging at 6-18 months after surgical resection. Ethical approval for this study was granted as a retrospective case note review by the Joint Research and Development Office at Great Ormond Street Hospital and UCL Great Ormond Street Institute of Child Health under 23NC04. Posterior fossa tumour patients were selected based on clinically-acquired diffusion MRI data, taken as standard clinical care. Patient IDs (i.e. PFXXX) were assigned as a part of this study and were not known outside the research group.

**Table 1.**
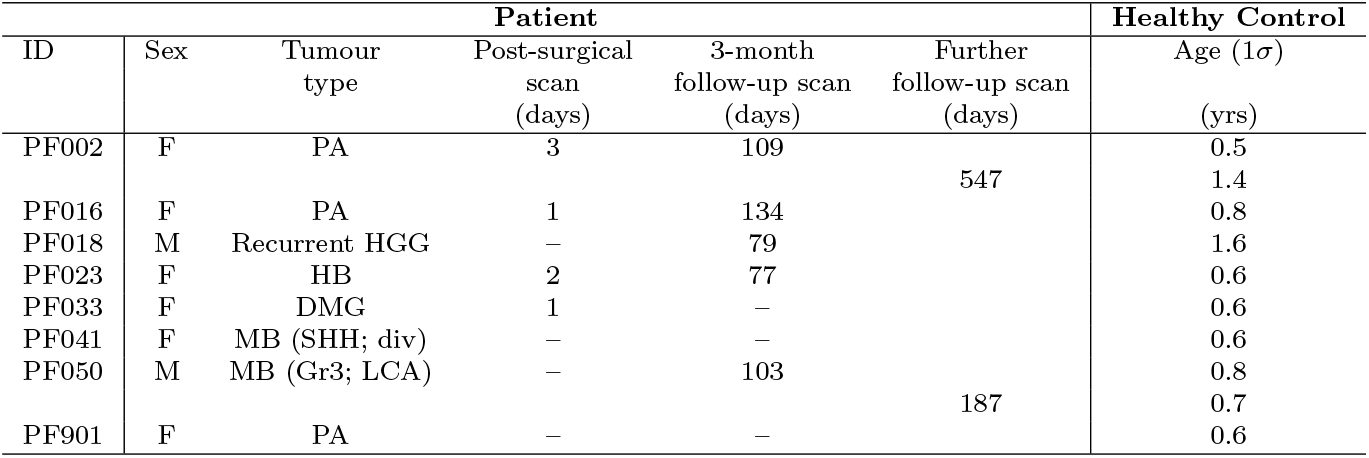
Patient and healthy control participant information. Patient age ranges were 7-15 years old at the time of the pre-surgical scan. Post-surgical, 3-month follow-up and further follow-up scan times are shown as the number of days after surgery. Tumour type acronyms are: medulloblastoma (MB), high grade glioma (HGG), diffuse midline glioma (DMG), pilocytic astrocytoma (PA), hemangioblastoma (HB). Medulloblastoma patients were molecular subgroups Group 3 (Gr3) or SHH and had histologies of divergent (div) or large cell/anaplastic (LCA). All patients had a group of 20 age- and sex-matched healthy controls. Controls were used for multiple patients when patients were similar in age. For the 6-18 month further follow-up scans (patients PF002 and PF050), the 20 age- and sex-matched controls were updated to reflect the increase in age compared to scans at earlier time points. The standard deviation (in years) around the control age mean is shown above, where the patient age fell within 1*σ* of the control group age mean.

**Table 2.**
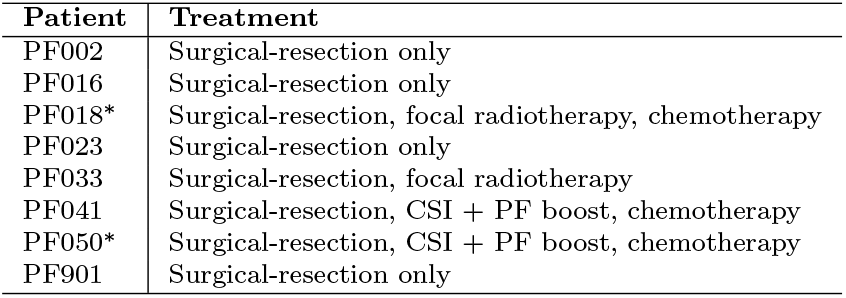
Treatment for the posterior fossa tumour patients. All patients had scans prior to any treatments, i.e. pre-surgery. Patients with ‘^*^’ show the patients with a scan after photon radiotherapy. Medulloblastoma patients are treated with craniospinal irradiation (CSI) with a boost to the posterior fossa (PF). Patient PF018 had previous treatment for HGG, which included focal radiotherapy administered 6 years prior to recurrence, followed by chemotherapy, with the official end of treatment approximately 4 years prior to recurrence. The treatment regime outlined above for PF018 was for the recurrence of HGG.

| Patient | Treatment |
| --- | --- |
| PF002 | Surgical-resection only |
| PF016 | Surgical-resection only |
| PF018* | Surgical-resection, focal radiotherapy, chemotherapy |
| PF023 | Surgical-resection only |
| PF033 | Surgical-resection, focal radiotherapy |
| PF041 | Surgical-resection, CSI + PF boost, chemotherapy |
| PF050* | Surgical-resection, CSI + PF boost, chemotherapy |
| PF901 | Surgical-resection only |

Healthy children were included in the analysis, for comparison to patients, originally recruited to existing ethically approved departmental studies, including UK Bromley Research Ethics Committee (REC 12/LO/0442), Research Ethics Service Committee London - Riverside (REC 14/LO/0115), London - West London & GTAC Research Ethics Committee (REC 15/LO/0347), London - London Bridge Research Ethics Committee (REC 16/LO/0256), UCL Institute of Education (REC approval number 1080), and London - Bloomsbury Research Ethics Committee (10/H0713/16). All control participants had no significant medical history and no radiological abnormalities were identified on MRI scans. In total, there were 102 control subjects (female-to-male ratio was 63:39; median age 10.1 years with age range 6.1-15.7 years).

### 2.2 MRI acquisition

MRI scans were performed at GOSH on a 3.0 T MAGNETOM Prisma (Siemens Healthcare) scanner using a 20 channel head receive coil. For control subjects, either a 64 or 20 channel head receive coil was used. The clinical MRI protocols for patients included multi-shell diffusion, an axial T2-weighted turbo spin-echo, fluid attenuated inversion recovery (FLAIR) sequences and pre- and post-gadolinium T1-weighted acquisitions. Volumetric T1-weighted images were acquired using an MPRAGE sequence with isometric 1.0 mm voxels, relaxation time (TR) =2300 ms and echo time (TE) =2.74 ms. The acquisition time was 5 minutes 21 seconds.

The multi-shell diffusion MRI sequence at our institution employed a diffusion-weighted spinecho single shot two-dimensional echo planar imaging acquisition, with multi-banded radio frequency pulses to accelerate volume coverage along the slice direction [32, 33]. There was a multiband factor of 2 employed over 66 slices of 2 mm thickness with 0.2 mm slice gap. The diffusion gradients were applied over 2 shells at b =1000 and 2200 s/mm^2^. There were 60 non-co-linear diffusion directions per shell with an additional 13 interleaved b_0_ (b =0 s/mm^2^; non-diffusion weighted) images. Imaging parameters were TR =3050 ms, TE =60 ms, field-of-view (FOV) =220*×*220 mm^2^, matrix size of 110*×*110, in-plane voxel resolution of 2.0*×*2.0 mm^2^, GRAPPA factor 2 and phase-encoding partial Fourier 6/8. Additionally, an identical b_0_ acquisition was performed to the diffusion-weighted scan but with the phase-encoding direction flipped by 180° in the anterior-posterior direction to correct artefacts related to susceptibility. The acquisition time was 7 minutes 50 seconds for the complete multi-shell diffusion sequence.

### 2.3 Image processing and modelling

Raw diffusion MRI files were processed using MRtrix3 [34] and Functional MRI of the Brain (FMRIB) Software Library (FSL v5.0) [35]. First, images were denoised using dwidenoise [36] employing a brain mask to improve processing speeds. A correction was made for Gibbs ringing artefacts using mrdegibbs [37]. Diffusion MRI and the reverse phase-encode b0 images were processed using dwifslpreproc, which uses the topup [38, 39] and eddy tools [40] from FSL to correct for susceptibility-induced distortions, artefacts from eddy currents and movement from the participant. Lastly, B1 field inhomogeneities were corrected using dwibiascorrect [39, 41]. The diffusion tensor was calculated per voxel using FSL’s dtifit. DTI parameter maps were computed for FA, MD, AD and RD.

The NODDI diffusion MRI modelling method was used to assess axon dispersion. This diffusion model assumes the tissue in a voxel is made of three compartments: an intracellular compartment (modelled by a set of sticks, representing the area bounded by the neurite membrane), an extra-cellular compartment (modelled by Gaussian anisotropic diffusion, representing the area around the neurites) and a compartment for free water or cerebrospinal fluid (CSF; modelled by isotropic Gaussian diffusion). This model estimates the density and coherence of neurites as well as the free water fraction or extent of CSF contamination. We were interested in the dispersion or coherence of the fibre tracts, known as the orientation dispersion index (ODI). Since NODDI is able to disentangle the effects of axon density from the dispersion of the axons, the ODI parameter was used to provide a more comprehensive understanding of white matter abnormalities also found using DTI.

Tissue segmentation was used to calculate the total brain volume and lateral ventricle volume of participants. Tissue segmentation was performed on T1-weighted MRI data using 5ttgen [42] script in MRTrix3, employing the FSL algorithm. This code segments a subject’s T1-weighted MR image into five tissue types, including grey matter, subcortical grey matter, white matter, CSF and pathological tissue.

### 2.4 Tract-based spatial statistics (TBSS)

TBSS [43] was employed to conduct a voxel-wise analysis of DTI parameters (FA, MD, AD and RD) and NODDI ODI. TBSS was used to (1) compare the posterior fossa tumour patient group at the pre-surgical time point to a group of age- and sex-matched healthy controls to assess statistical differences in DTI parameters and (2) use a ‘one-against-many’ approach to compare each individual patient to 20 age- and sex-matched healthy controls to assess statistical differences in DTI and ODI parameters at the different time points (pre-surgery, post-surgery, 3-month follow-up and any further follow-up). For the one-against-many approach, the patient age was within 1*σ* of the control group mean age (details for the standard deviation of control group ages can be found in Table 1). In the TBSS analysis, the patient and controls were aligned into the Montreal Neurological Institute (MNI152) common space using FMRIB’s Nonlinear Image Registration Tool (FNIRT). A mean FA image was generated from the individual patient and control group images (thresholded at FA of 0.2), which was used to generate a FA ‘skeleton’ of white matter tracts that is study-specific, which represents the centre of the white matter tracts throughout the brain. Each patient and control image was aligned to the FA skeleton, which allowed for the voxel-wise comparison of an individual patient to their control group. The FSL randomise [44] function was used for voxel-wise statistics on the skeleton of the DTI parameter maps, which used the threshold free cluster enhancement option and family-wise error corrected at p *<*0.05.

To assess the quality of TBSS results and ensure significant differences in DTI parameters in the white matter skeleton between the patient and control group corresponded to voxels in the white matter tracts, the results were transformed back into the patient’s native space. A mask was generated to include the supratentorial region of the brain and exclude spurious signal that corresponded to ventricle space (additional methods are available as supplementary material accompanying the online article). The JHU ICBM-DTI-81 white matter labels atlas [45], consisting of 48 distinct white matter tracts, was used to identify clusters along the white matter skeleton with significantly different DTI parameters from the TBSS analysis.

### 2.5 Lateral ventricle volumes

To assess the degree of patient obstructive hydrocephalus due to the presence of a posterior fossa tumour, lateral ventricle volumes were calculated from the tissue-segmented T1-weighted images (described in Section 2.3). This method used a binary mask to extract the lateral ventricles from CSF images. The binary mask template was originally produced for the Automatic Lateral Ventricle delIneatioN (ALVIN) method [46], based on an average of 275 CSF images from healthy subjects aged 18 to 94. The mask included the entire lateral ventricular system as well as the temporal horns. First, the mask was qualitatively assessed by applying the mask on averaged CSF maps of the patients and the controls to ensure it covered the lateral ventricles. The mask was then applied to individual patient and control CSF images and manually corrected for boundary changes when required. Lateral ventricle volumes were then calculated in mm^3^ by summing the intensity over the spatially normalised CSF segmented image. Third ventricle volumes were not calculated as it is not surrounded by white matter; the fourth ventricle was obscured by the tumour in almost all cases and was also not evaluated.

## 3 Results

### 3.1 Group analysis

The group of 8 posterior fossa tumour patients were compared to a group of healthy controls at the pre-surgical time point. The control group was representative of the patients, where 3 age- and sex-matched controls were chosen to match each patient. Therefore, the pre-surgical patient group was compared to a group of 24 controls using TBSS comparing DTI parameters. The group of 24 controls had an age range of 7.3-14.8 years (mean age of 10.4 years) and the female-to-male ratio was 18:6 (75% female). The patient group at the pre-surgical time point had an age range of 7.5-14.6 years (mean age of 10.4 years) and the female-to-male ratio was 6:2 (75% female).

DTI parameters FA, MD, AD and RD were significantly different (p-value *<* 0.05) between the patient and control group in many white matter tracts throughout the brain pre-surgery (Figure 1). There was widespread decreased FA and increased MD, AD and RD in regions corresponding to the corpus callosum (body, splenium and genu), fornix, corona radiata and posterior thalamic radiations. The FA decrease corresponded to 36.1% of the white matter skeleton. Diffusivity increases corresponded to 18.6% (MD), 16.4% (AD) and 35.9% (RD) of the white matter skeleton.

**Figure 1.**
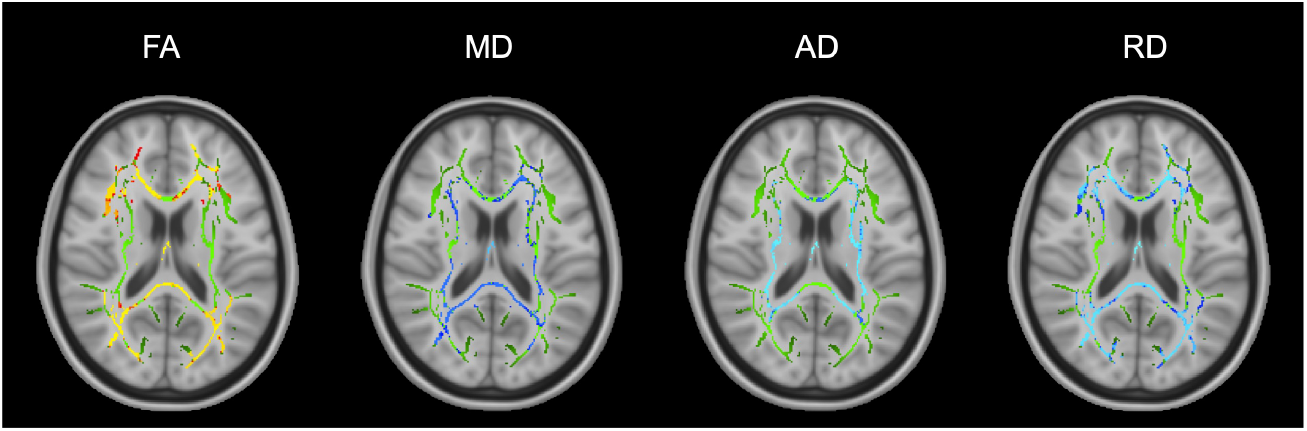
TBSS results for the group of all 8 posterior fossa tumour patients at the pre-surgical time point. Green denotes the white matter skeleton where voxels are not significantly different between the patient and control groups. The yellow to red colour scale denotes voxels that are significantly decreased in the patient group and light blue to blue colour scale denotes voxels that are significantly increased in the patient group. Significant p-values for FA are *<* 0.05 (yellow values are *<* 0.01). Significant p-values for MD are *<* 0.05 (light blue values are *<* 0.015). AD p-values are *<* 0.03 (light blue values are *<* 0.005). RD p-values are *<* 0.05 (light blue values are *<* 0.005).

### 3.2 Individual analysis

#### 3.2.1 Pre-surgical time point

**DTI** Out of the 8 patients individually compared to 20 age- and sex-matched controls, 5 patients exhibited significant changes in FA prior to surgery, including 3 patients with widespread changes and a further 2 patients with localised changes. For patients with widespread changes in FA, more than 5 % of the total white matter skeleton and *>* 15% of the labelled white matter tracts identified in the JHU ICBM-DTI-81 atlas.

The pattern of widespread FA changes can be split into two categories:

1. An increase in FA corresponding to the periventricular white matter (PVWM), i.e. internal capsule and corona radiata (patients PF002 and PF050, Figures 2A, 2C).
2. A decrease in FA (patient PF016, Figure 2B) in all white matter tracts.

**Figure 2.**
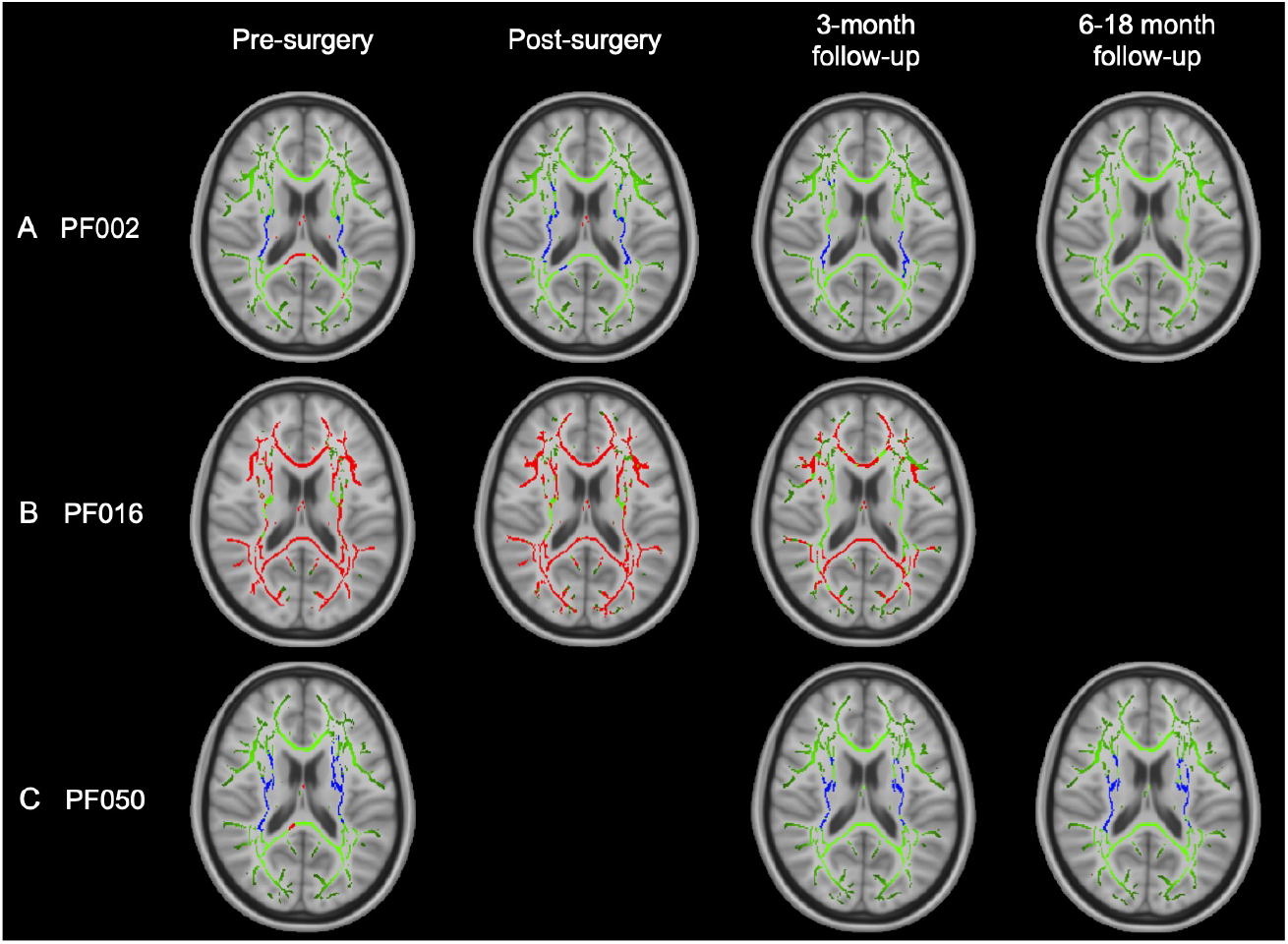
TBSS results for individual patients with widespread, significant changes in FA, including pilocytic astrocytoma patients PF002 (A) and PF016 (B) and medulloblastoma patient PF050 (C). Green corresponds to the white matter skeleton where voxels are not significantly different between the individual patient and group of controls. Red voxels correspond to areas of the white matter skeleton that are significantly decreased in the patient (p-value *<* 0.05), and blue voxels are significantly increased in the patient (p-value *<* 0.05). From left to right, the image timeline is from pre-surgery, post-surgery, follow-up at 3 months and a further follow-up at 6-12 months (as in Table 1).

These patients also had significant changes in diffusivity throughout the white matter skeleton (details of MD, AD and RD are available as supplementary results accompanying the online article). Diffusivity changes were more complex than FA:

1. PF002: MD, AD and RD (Figure S3) showed a pattern of increased values near the ventricles, surrounded by white matter with decreased values. In PVWM, specifically the internal capsule and corona radiata, both MD and RD were decreased and there were areas of both increased and decreased AD. The posterior thalamic radiations exhibited decreased FA, MD and AD with increased RD. The fornix was the only area to have decreased FA with increased MD, AD and RD.
2. PF016: There were widespread increases in MD and RD (Figure S6). AD showed a more complex pattern similar to PF002 with increased values in PVWM, surrounded by decreased values distal to the ventricles.
3. PF050: AD was primarily increased, particularly in PVWM, corresponding to the internal capsule and corona radiata, while RD was decreased (Figure S9). MD was mainly unchanged. The fornix deviated from the other areas of white matter with decreased FA and increased MD, AD and RD.

Furthermore, there were 2 patients with localised decreases in FA at the pre-surgical time point, and there were no changes in diffusivity parameters for these patients. PF018 had decreased FA in the body and genu of the corpus callosum and the corona radiata (1.2% of the total white matter skeleton) and PF901 had decreased FA in the left corona radiata (0.3% of the total white matter skeleton). However, patient PF018 had recurrent HGG and had previously received focal radiotherapy, so the pre-surgical treatment for this patient could have been due to earlier treatment.

### NODDI ODI

Prior to surgery, only patients PF002, PF016 and PF050 with widespread FA changes across the white matter skeleton were found to have significant alterations in ODI compared to their control groups (Figure 3). These changes were found in two patterns:

1. A widespread decrease in ODI corresponding to PVWM, i.e. internal capsule and corona radiata (patients PF002 and PF016), surrounded by an increase in ODI.
2. A widespread decrease in ODI in PVWM only (patient PF050).

**Figure 3.**
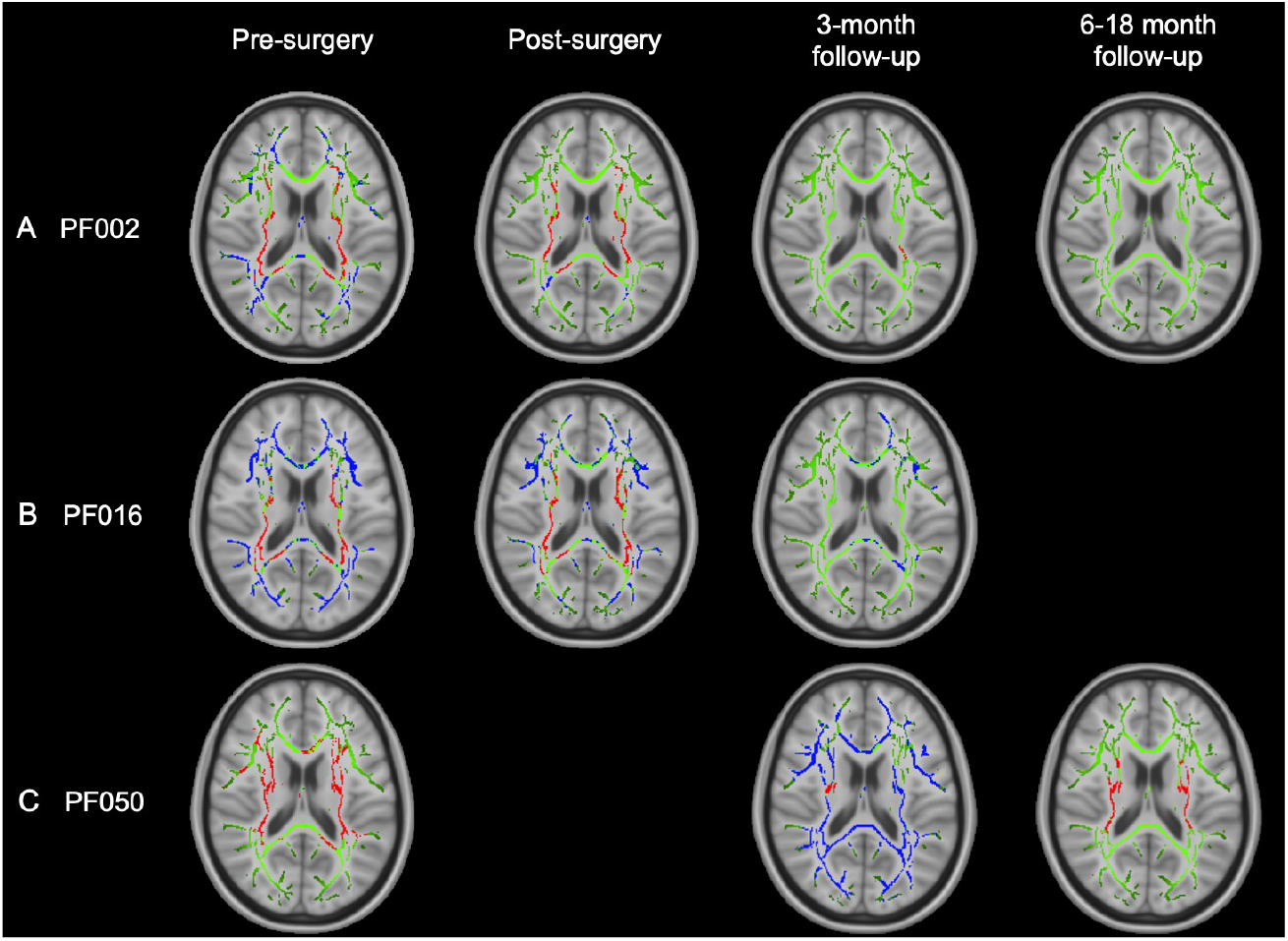
TBSS results for individual patients with widespread, significant changes in ODI, including pilocytic astrocytoma patients PF002 (A) and PF016 (B) and medulloblastoma patient PF050 (C). Green corresponds to the white matter skeleton where voxels are not significantly different between the individual patient and group of controls. Red voxels correspond to areas of the white matter skeleton that are significantly decreased in the patient (p-value *<* 0.05), and blue voxels are significantly increased in the patient (p-value *<* 0.05). From left to right, the image timeline is from pre-surgery, post-surgery, follow-up at 3 months and a further follow-up at 6-12 months (as in Table 1).

#### 3.2.2 Longitudinal changes

**DTI** Over time, DTI parameters did not fully return to normal in patients with pre-surgical changes (Figure 2). At the post-surgical time point, patients PF002, PF016 and PF050 generally had DTI parameters that were relatively similar or potentially improved compared to their pre-surgical scan. At follow-up, there was evidence of improvement but results were mixed:

1. PF002: FA returned to normal values over an 18 month period after surgery, specifically in PVWM (internal capsule and corona radiata), which originally had elevated FA. However, there was a decrease in diffusivity (MD, AD and RD) that became more widespread through the white matter skeleton at 3 months and 18 months after surgery. More details can be found in the supplementary results (Figures S3-S5).
2. PF016: FA remained significantly decreased across many white matter tracts from the pre-surgical time point to the 3 month follow-up. Diffusivity parameters mainly returned to normal values. AD was only increased in the fornix and RD was increased in the fornix, body and splenium of the corpus callosum and areas corresponding to PVWM (internal capsule and corona radiata). More details can be found in the supplementary results (Figures S6-S8).
3. PF050: At 3 months post-surgery (after radiotherapy), FA remained significantly increased in PVWM, corresponding to the internal capsule, corona radiata and external capsule. The only change in diffusivity corresponded to increased RD in the fornix, which also showed decreased FA. At 6 months post-surgery (3 months after CSI), MD was decreased in the left hemisphere. RD was decreased in PVWM, similar to the pre-surgical scan. More details can be found in the supplementary results (Figures S9-S11).

In addition to the 3 patients with changes at the pre-surgical time point, there were 2 other patients, PF018 and PF033, with decreased FA post-surgery. In patient PF018, there was a change in the number of voxels with decreased FA between the pre-surgical time point and 3 month follow-up, increasing from 1.2% to 10.8% of the white matter skeleton. This corresponded to the corpus callosum (genu, body and splenium), corona radiata and posterior thalamic radiations. Patient PF033 was found to have decreased FA in 15.9% of the voxels in the post-surgical scan, which corresponded to the corpus callosum (genu, body and splenium), internal capsule, fornix, corona radiata, posterior thalamic radiations, superior longitudinal fasciculus, inferior fronto-occipital fasciculus and uncinate fasciculus.

### NODDI ODI

After surgery, only patients with pre-surgical changes in ODI were found to have longitudinal changes in this parameter, including PF002, PF016 and PF050. Over time, patients PF002 and PF016 had ODI parameters that began to return to normal after 3-18 months. A different trend was observed in patient PF050. Initially PF050 had decreased ODI in PVWM. In the 3 month follow-up (after radiotherapy), there was widespread increase in ODI in all white matter tracts. At the 6 month follow-up scan (3 months post-radiotherapy), there was a decrease in ODI in PVWM that resembled the pre-surgical TBSS analysis.

#### 3.2.3 Lateral ventricle volumes

All patients had enlarged lateral ventricles (p-value*<*0.05) when compared to their age- and sex-matched controls (Figure 4). The 3 patients with the most enlarged lateral ventricles (lateral ventricle volumes *>*4% of the total brain volume) correspond to the 3 patients with the most widespread changes in FA.

**Figure 4.**
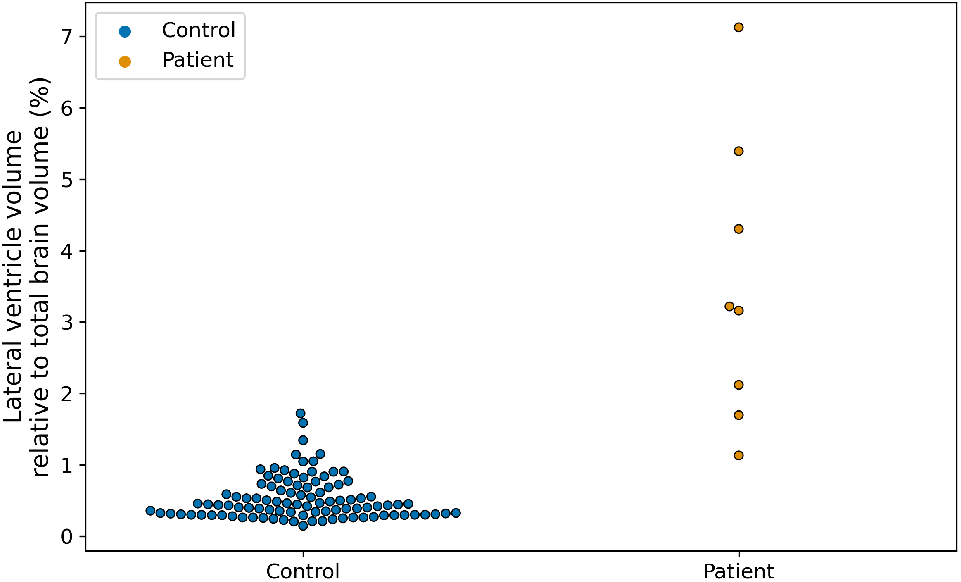
Comparison of the lateral ventricle volumes relative to the total brain volume (%) of the patients at the pre-surgical time point and all controls.

Over time, the lateral ventricles decreased in size for most of the patients following surgery (PF002, PF016, PF023, PF050), but remained significantly enlarged for patients PF002, PF016 and PF050 compared to their control groups. In patients PF018 and PF033, the ventricles grew larger after surgery (PF018: 1.1% to 1.8%; PF033: 1.7% to 1.9% of the total brain volume).

## 4 Discussion

Our study is the first to demonstrate, using diffusion imaging, significant changes to supratentorial white matter caused by the presence of an infratentorial tumour prior to treatment. Using a one-against-many comparison, we found that observed white matter abnormalities are complex and patient-dependent, indicating that white matter changes may be missed using analysis techniques that group posterior fossa brain tumour patients together for comparison with a control group.

In the posterior fossa patient group analysis of pre-surgical images, we found widespread decreased FA and increased diffusivity (MD, AD and RD) throughout numerous white matter tracts compared to the healthy control group. At this time point, all patients were found to have significantly enlarged lateral ventricles relative to total brain volume, likely due to hydrocephalus caused by the tumour obstructing the fourth ventricle. These DTI abnormalities could indicate that there was increased diffusion in patient white matter and the diffusion was less directional (more isotropic). This was consistent with oedema or damage to white matter, e.g. reduced coherence of white matter tracts, decreased fibre density or damage to white matter integrity [47, 48]. Obstructive hydrocephalus can cause a rupture of the ventricular ependymal lining due to increasing intraventricular pressure. CSF can then move into the extracellular spaces leading to increased interstitial fluid, i.e. oedema [49, 50].

Even though white matter abnormalities were present in the pre-surgical group analysis, the one-against-many technique was able to highlight the heterogeneity of white matter abnormalities in individual posterior fossa tumour patients, which were not captured when comparing the patient group to the control group. Prior to surgery, we found that 5 out of 8 patients had evidence of supratentorial white matter injury using the one-against-many approach. Three of the patients, including PF002, PF016, PF050, showed widespread changes in FA in *>*5% of the white matter skeleton. These patients had the most enlarged ventricle volumes at the pre-surgical time point, corresponding to lateral ventricles *>*4 % of the total brain volume (*>*10*σ* above healthy control mean). Other patients with DTI changes prior to surgery included PF018, a high-grade glioma patient, and PF901, a pilocytic astrocytoma patient. Both patients had decreased FA in small, localised areas with no changes in diffusivity. While these patients had enlarged ventricles, the degree of ventricular enlargement was not as extreme as the patients with widespread DTI changes. This finding suggests that hydrocephalus potentially impacts the brain white matter microstructure prior to surgery.

Compression effects due to hydrocephalus were also seen in the analysis of the axonal dispersion using NODDI ODI. In patients PF002, PF016 and PF050, the pattern of decreased ODI in PVWM prior to surgery may be an indicator of tissue compression caused by enlarged ventricles, where we expect compression to cause a decrease in axon dispersion (or an increase in tract coherence) in PVWM. In patients PF002 and PF016, the area surrounding PVWM was seen to have decreased coherence (increased ODI), which is potentially related to the greater size of the ventricles.

The widespread white matter changes found before surgery were observed to persist over time, particularly FA. The widespread pre-surgical changes in FA were observed in different patterns, including a decrease in FA throughout supratentorial white matter (PF016: pilocytic astrocytoma) and an increase in FA corresponding to periventricular white matter (PF002: pilocytic astrocytoma; PF050: medulloblastoma). These FA patterns are consistent with studies that have used DTI to understand white matter microstructure in patients with hydrocephalus (e.g. Assaf et al. [51], Ben-Sira et al. [52], Air et al. [53], Hattingen et al. [54], Osuka et al. [55], Yuan et al. [56]). Increased FA in the periventricular region may be the result of the compression of these white matter tracts due to enlarged lateral ventricles [51, 57, 58], where fibres that are homogeneously aligned will be packed together more closely. Decreased FA may be indicative of microstructural white matter damage or diffuse oedema caused by increased pressure in the ventricles that disrupts the ventricular ependymal lining. Chronic tissue compression from hydrocephalus has previously been found to result in tissue degeneration, including demyelination, axonal loss, gliotic change, loss of neuronal transmission and higher packing of white matter fibres [59–61], and there is evidence that the largest ventricles lead to worse PVWM destruction and deficits in motor and cognitive function [62]. This could lead to cumulative damage to white matter, for example, when adjuvant treatment involves radiotherapy. To improve outcomes of patients, pre-treatment imaging could be considered during individual treatment planning and risk evaluation to avoid further injury to white matter or to minimise the volume of white matter damage.

In addition to studying the effects of hydrocephalus, NODDI may be beneficial for interpreting short and long-term white matter abnormalities due to radiotherapy. For example, the medulloblastoma patient PF050 was treated with surgical resection, CSI with a posterior fossa boost and chemotherapy. While the the TBSS analysis of the ODI parameter originally showed evidence of increased white matter coherence (decreased ODI) in PVWM prior to surgery, at the 3-month follow-up scan there was evidence of widespread decreased coherence (increased ODI) across all white matter tracts. This time point corresponded to the end of radiotherapy. Decreased coherence in white matter can indicate axonal disorganisation or increased extra-axonal space [63]. This is consistent with early radiotherapy effects, such as inflammation, which is expected to contribute to demyelination over time caused by the apoptosis of oligodendrocytes [64]. At the 6-month follow-up scan (3 months after treatment), the TBSS analysis of ODI returned to a similar pattern as the pre-surgical analysis (i.e. decreased dispersion at PVWM). There was also new evidence of white matter abnormalities in the DTI analysis at this time point, indicated by decreased MD in the left hemisphere white matter tracts (Figure S9).

The one-against-many technique is a novel approach in diffusion MRI, enabling a more individualised understanding of how white matter integrity is impacted over time in paediatric brain tumour patients. A benefit of the one-against-many approach is that it allows for the variability between white matter abnormalities in patients of similar ages and with the same tumour type to be explored. For example, we compare the female pilocytic astrocytoma patients PF002, PF016 and PF901. These patients had no further treatment after surgical resection of the tumour. The patients showed different white matter abnormalities, likely due to the effects of hydrocephalus, evidenced by an increase in FA in PF002 in PVWM, a decrease in FA in PF016 and both patients had evidence of compression effects in ODI (increased coherence in PVWM surrounded by decreased coherence further from the ventricles). PF901 was found to have a localised decrease in FA in the left corona radiata and no change in ODI or diffusivity. The differences between these patients are likely due to the ventricle sizes, where PF016 had the most enlarged ventricles compared to total brain volume (7.1%) compared to PF002 (5.4%) and PF901 (3.2%). Over time, both PF002 and PF016 have FA and ODI values that began to return to normal over a 3-18-month period, but there were differences in how diffusivity parameters changed over time. MD values across the white matter skeleton showed large-scale significant decreases compared to control values that affected more of the white matter skeleton in the later timescales 3-18 months after surgery, which could be due to gliosis (Figure S6). This may indicate that the effects of hydrocephalus on PF002 are different and more long-lasting than PF016 or PF901.

The ability to conduct the one-against-many analysis quickly - approximately 13 minutes for FA and an additional 6 minutes for other diffusivity parameters - makes it feasible to incorporate into clinical workflows. This has important implications confirming that brain injury can occur pre-treatment and distant to the tumour location as evidence by the abnormal supratentorial white matter seen in childhood posterior fossa tumours. This may allow tailoring of future therapeutic or rehabilitation strategies, as white matter abnormalities detected early could inform adjustments in treatment plans to mitigate long-term neurocognitive deficits. Cognitive follow-up will be essential in future studies to correlate pre-treatment white matter changes with cognitive outcomes. Future work could (1) explore the differences in microstructure and motor and cognitive function in patients with infratentorial tumours to better understand the impact of obstructive hydrocephalus on long-term outcomes, (2) understand risk factors for white matter injury that persists despite effective treatment of ventriculomegaly and (3) identify the contribution of adjuvant therapy for individual patients.

Our study has several limitations. The sample size of this study is small, though it is comparable to other exploratory studies involving the use of DTI to understand microstructural changes in white matter in posterior fossa tumour patients at post-surgical and/or post-treatment time periods. With larger patient numbers, the underlying cause of DTI changes prior to treatment could be further investigated. Furthermore, cumulative effects of the tumour and treatments, which may lead to worse neurocognitve outcomes, could be explored. Additionally, one of the challenges of a one-against-many approach is the need to acquire at least 20 age- and sex-matched healthy controls for each patient on the same MRI machine. The control group for each of the patients in this work are not evenly spread in age, though care was taken in ensuring the age of the patient was within 1*σ* of the mean control age. Discrepancies in the spread of ages in the control group for each patient could lead to the TBSS analysis being more or less sensitive depending on if there was control data that better matched the age of a patient.

## 5 Conclusions

This study is the first to demonstrate the presence of white matter abnormalities prior to treatment in posterior fossa brain tumour patients using diffusion MRI. This knowledge may contribute to disentangling the causes of white matter damage in the brain, either from the tumour, associated hydrocephalus or from treatments such as radiotherapy. Furthermore, the one-against-many approach shows that pre-treatment white matter changes are patient-dependent, which may have a bearing on the ultimate treatment response and neurocognitive outcomes of patients. This approach underlines the variation and individuality of white matter injury in children with a posterior fossa tumour, and has the potential to define risk factors for irreversibility despite post-treatment reduction in ventricular size. This may have implications for understanding white matter vulnerability in individual children, tailoring of adjuvant therapy to reduce risk and early planning of rehabilitation strategies. These white matter abnormalities may, therefore, need to be considered in treatment planning for individual patients to improve outcomes and prevent cumulative white matter damage.

## Data Availability

All data produced in the present study are available upon reasonable request to the authors.

## Abbreviations

AD: axial diffusivity;
CSI: craniospinal irradiation;
CSF: cerebral spinal fluid;
DMG: diffuse midline glioma;
DTI: diffusion tensor imaging;
FA: fractional anisotropy;
GOSH: Great Ormond Street Hospital for Children;
HB: hemangioblastoma;
HGG: high-grade glioma;
MB: medulloblastoma;
MD: mean diffusivity;
MRI: magnetic resonance imaging;
NODDI: Neurite Orientation Dispersion Diffusion Imaging;
ODI: orientation dispersion index;
PA: pilocytic astrocytoma;
PVWM: periventricular white matter;
RD: radial diffusivity;
TBSS: tract-based spatial statistics.

## Acknowledgments

EDM is funded by the UCL EPSRC CDT i4health (grant reference EP/ S021930/1), and EDM and DRH are funded by the NIHR Great Ormond Street Hospital Biomedical Research Centre (NIHR-INF-2647). JAD is supported by the Radiation Research Unit at the Cancer Research UK City of London Centre Award [C7893/A28990].

## Financial disclosure

None reported.

## Conflict of interest

The authors declare no potential conflict of interests.

## Supporting information

Additional supporting information may be found in the online version of the article, including supplemental methods (Figures S1 and S2) and results (Figures S3-S11).

## Notes

### Competing Interest Statement

The authors have declared no competing interest.

### Author Declarations

Ethical approval for this study was granted as a retrospective case note review by the Joint Research and Development Office at Great Ormond Street Hospital and UCL Great Ormond Street Institute of Child Health under 23NC04. Healthy children were included in the analysis, for comparison to patients, originally recruited to existing ethically approved departmental studies, including UK Bromley Research Ethics Committee (REC 12/LO/0442), Research Ethics Service Committee London - Riverside (REC 14/LO/0115), London - West London & GTAC Research Ethics Committee (REC 15/LO/0347), London - London Bridge Research Ethics Committee (REC 16/LO/0256), UCL Institute of Education (REC approval number 1080), and London - Bloomsbury Research Ethics Committee (10/H0713/16).

