## Supplementary Materials for "Evidence of supratentorial white matter injury prior to treatment in children with posterior fossa tumours using diffusion MRI"

Supplementary materials for *Evidence of pre-treatment abnormal supratentorial white matter in children with posterior fossa tumours from diffusion MRI*

### Supplemental methods

#### Supratentorial mask

After the TBSS analysis was performed (Section 2.4), FA values in voxels that were identified as significantly different between the patient and control group were transformed back into the patient's native space to ensure voxels were originating from white matter tracts. This was done using FSL's `deproject`. In Figure S1 A, there is an example from PF002 of FA values that were significantly increased (blue) and decreased (red) in the nonlinearly registered space for the patient (aligned into the MNI152 common space). It can be seen that some of the voxels with significantly decreased FA corresponded to spurious signal taken from the ventricle space as opposed to white matter tracts. A mask was designed (Figure S1 B), shown on the MNI T1 template in yellow, to only include the supratentorial region of the brain and exclude any spurious signal from the lateral ventricles.

After applying the mask to the TBSS results, only the signal from white matter tracts in the supratentorial region was kept for the analysis of white matter. Figure S2 shows PF002 TBSS results of FA after the application of the mask. TBSS results can be seen on the nonlinearly registered space for PF002 (A) and on the MNI T1 template (B). The supratentorial mask was applied to TBSS results for all patients.

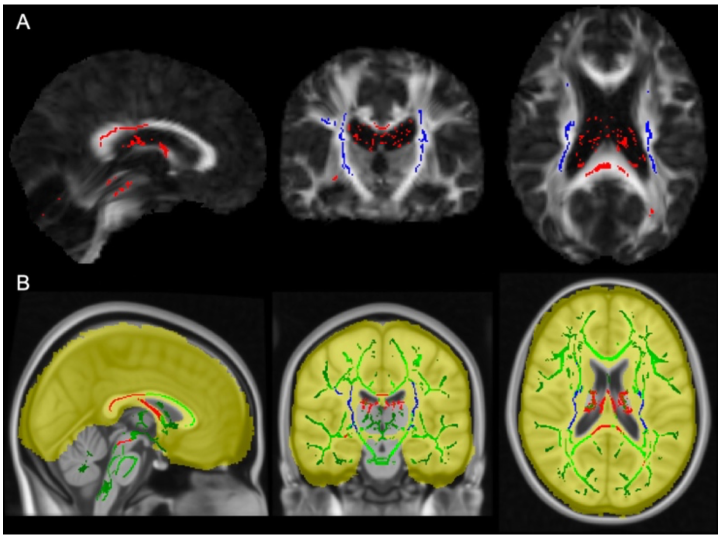


***Figure S1:*** Example of mask generation. (A) TBSS results of increased and decreased FA on the nonlinearly registered space for PF002. (B) Yellow corresponds to the design of the supratentorial mask that excluded spurious voxels in between the lateral ventricles.

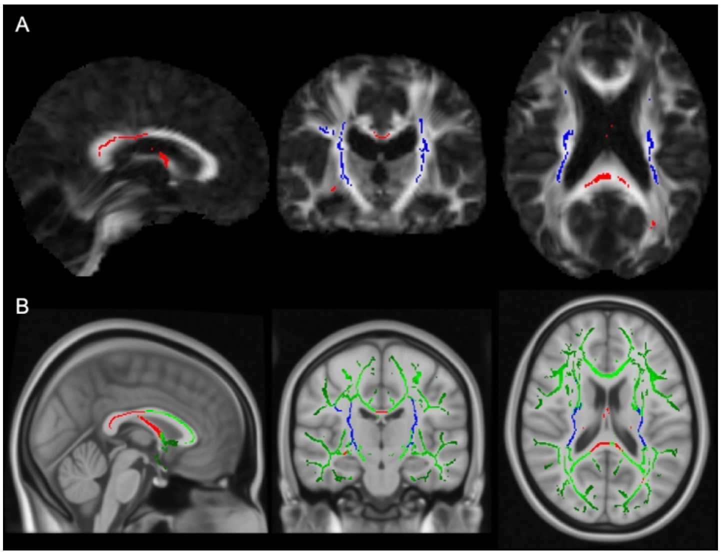


***Figure S2:*** Examples of the TBSS results after the application of the supratentorial mask. (A) TBSS results of significant FA changes on the nonlinearly registered space for PF002. (B) Significant FA changes on the template space for PF002 after the application of the mask.

**Supplemental results: DTI analysis**

Patient PF002:

Structural and diffusion MR imaging was taken prior to surgery, 3 days post-surgery, 3 months after surgery and 18 months after surgery. There was no further treatment after surgical resection of the tumour. DTI results for FA can be found in Figure 2 of the main text and diffusivity parameters (MD, AD and RD) in Figure S3.

Results from the TBSS analysis using the one-against-many technique suggested complex alterations in supratentorial white matter. The FA in PVWM was originally significantly increased in the patient, likely from the enlarged lateral ventricles due to obstructive hydrocephalus caused by the tumour. FA values returned to normal levels over the following 12 months after surgery. Figure S4 shows median FA values in periventricular white matter for the patient and the control group at each time point. In this region, median FA values were elevated in the patient compared to the control group prior to surgery (22.8%), 3 days after surgery (29.2%) and 3 months after surgery (21.8%). At the 18 month follow-up scan, FA in PVWM in the patient was only elevated by 9.8% compared to the control group and was no longer significantly increased at the voxel level in the TBSS analysis.

TBSS results from diffusivity indicated there may be microstructural abnormalities that cannot be seen using FA or NODDI ODI. Similar to FA, AD was increased in PVWM prior to surgery before returning to normal after surgery, which was consistent with compression due to enlarged ventricles from the infratentorial tumour. However, there were widespread regions of increased MD and AD in white matter more distal from the ventricles that became more widespread through the white matter skeleton at 3 months and 18 months after surgery. At the pre- and post-surgical time points, 23.5% and 15.8% of the total voxels in the white matter skeleton had significantly decreased MD, respectively. This expanded to 66.5% and 58.6% of the total white matter skeleton at 3 months and 18 months post-surgery.

Figure S5 shows MD values across supratentorial white matter tracts in the patient and control group. The median MD values were decreased in the patient before surgery (2.5%) and after surgery (3.1%). MD was decreased across more white matter tracts at the 3 month follow-up (5.5% decreased compared to controls) and at the 18 month follow-up (5.3%).

In general, a reduction in diffusivity is thought to be an indicator of increased cellular density (Alexander et al. 2007), which could be due to gliosis caused by the initial obstructive hydrocephalus (e.g. Del Bigio 1993, Del Bigio et al. 2003). Decreased diffusivity has also been observed in ischaemic events (van Gelderen et al. 1994). However, MD is found to normalise several days after the initial ischaemic event before increasing due to cerebral softening.

Patient PF016:

Structural and diffusion MR imaging were taken prior to surgery, 3 days post-surgery and 3 months after surgery. There was no further treatment after surgical resection of the tumour. DTI results for FA can be found in Figure 2 in the main text and diffusivity parameters (MD, AD and RD) in Figure S6.

Results from the TBSS analysis using the one-against-many technique suggested widespread alterations in supratentorial white matter. FA throughout white matter tracts was decreased initially at the pre-surgical time point, affecting 76.9% of the white matter skeleton, potentially due to ventricular enlargement and/or diffuse oedema, where PF016 had the largest lateral ventricles compared to total brain volume out of all patients and controls. FA values do not fully return to normal levels over the next 3 months after surgery, where 34.9% of the white matter skeleton at the 3 month follow-up continued to have decreased FA.

Figure S7 shows median FA values across supratentorial white matter tracts for the patient and control group at each time point. Median FA values of PF016 were decreased by 31.1% and 31.4%

before and directly after surgery compared to control group median values. At the 3 month follow-up scan, the FA median value was 15.8% below the control group, which was consistent with some supratentorial white matter recovery.

Diffusivity results were consistent with white matter recovery over time, where initially increased MD, AD and RD returned to more normal values across the majority of the white matter tract (Figure S6). Figure S8 shows MD values across supratentorial WM tracts in the patient and control group. The median MD values in PF016 were increased by 11.2% and 11.5% compared to the control group median at the pre- and post-surgical time points. At the 3 month follow-up scan, the median MD was 3.9% elevated compared to the control group.

Patient PF050:

Structural and diffusion MR imaging were taken prior to surgery, 3 months post-surgery (directly after radiotherapy) and 6 months after surgery. Treatment included surgical resection, CSI with posterior fossa boost and chemotherapy. DTI results for FA can be found in Figure 2 in the main text and diffusivity parameters (MD, AD and RD) in Figure S9.

Results from the TBSS analysis using the one-against-many technique suggested alterations primarily in PVWM. FA in this region was increased at the pre-surgical time point, potentially due to compression from ventricular enlargement (i.e. hydrocephalus). FA values do not fully return to normal levels over the next 6 months after surgery (3.3% of the total white matter skeleton). Figure S10 shows median FA values across the PVWM for the patient and control group at each time point. Median FA values of PF050 were increased by 23.0% prior to surgery and 17.9% and 16.8% at the 3 month and 6 month follow-up scans compared to control group median values. This demonstrated that abnormalities in white matter caused by secondary complications, such as hydrocephalus, can last for several months after resection of infratentorial tumours, including throughout further treatments (e.g. radiotherapy and chemotherapy).

Diffusivity results are less clear (Figure S9). The increase in AD and decrease in RD in periventricular white matter prior to surgery was consistent with compression effects from ventricular enlargement. This appeared to return to normal at the 3 month follow-up scan (directly after the end of radiotherapy). However, there was a decrease in MD noted in the left corona radiata (7.9% of the white matter skeleton) and decrease in RD in PVWM (2.0% of the white matter skeleton) at the 6 month follow-up scan.

Figure S11 shows MD values across the supratentorial white matter skeleton in PF050 and the control group. From the TBSS analysis, MD did not significantly differ between the patient and control group on a voxel level until the 6 month follow-up scan. However, median MD values in PF050 were decreased by 1.5% and 1.8% at the pre-surgical and 3 month follow-up time points. The median MD value continued to decrease by 4.2% across all white matter tracts by the 6-month follow-up scan, which could indicate MD is an indicator of radiation effects.

### Patient PF002

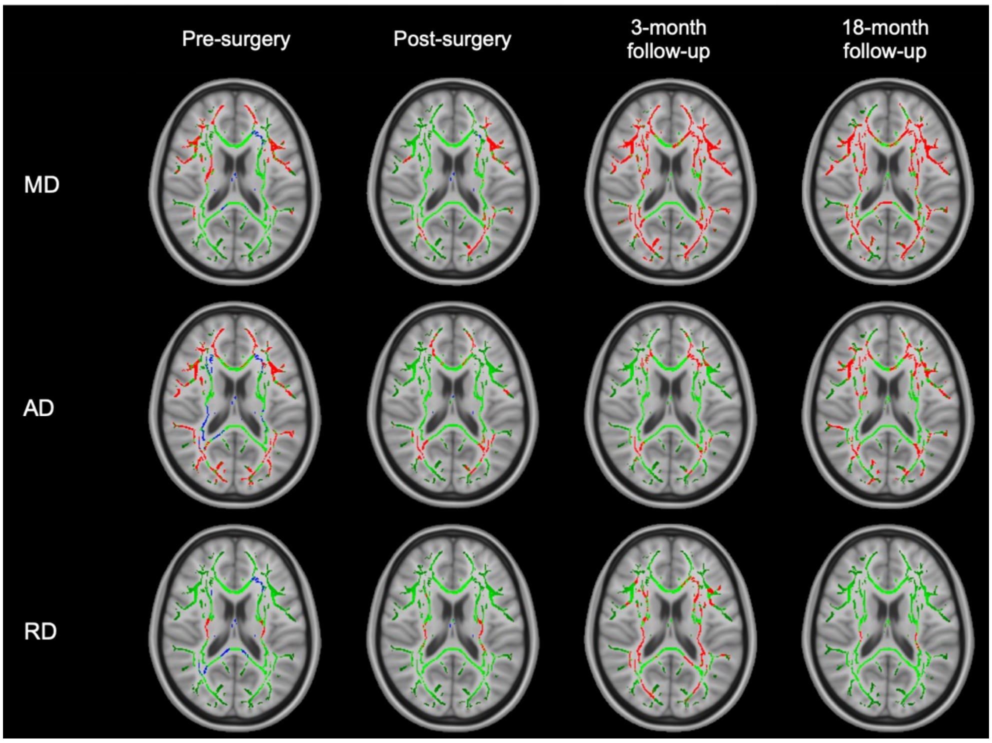


***Figure S3:*** Patient PF002 TBSS results of DTI diffusivity parameters (MD, AD and RD). Green voxels correspond to the white matter skeleton, where voxels were not significantly different between the individual patient and group of controls. Red voxels correspond to areas of the white matter skeleton that were significantly decreased in the patient (p-value < 0.05), and blue voxels were significantly increased in the patient (p-value < 0.05).

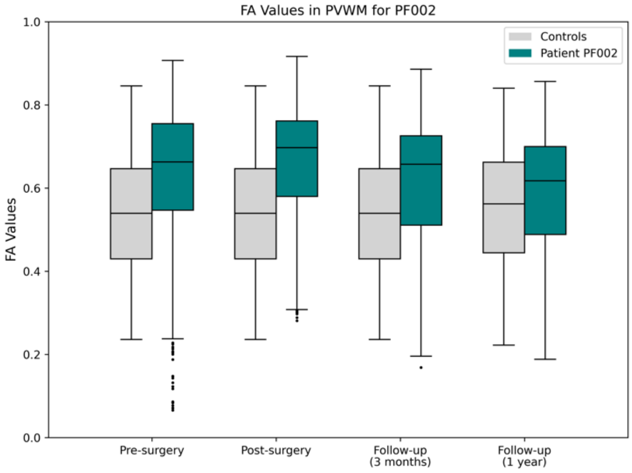


***Figure S4:*** Median FA values from PVWM in patient PF002 compared to the average median FA values from PVWM in the control group.

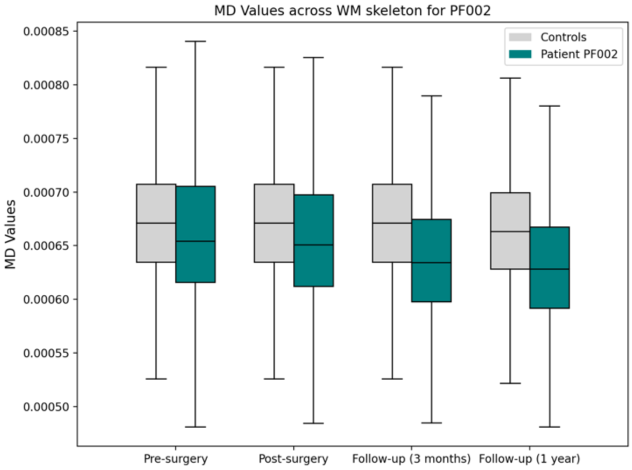


***Figure S5:*** Median MD values across the full white matter skeleton in patient PF002 compared to the average median MD values across the full white matter skeleton in the control group. Outliers have been removed from the box plot for clarity.

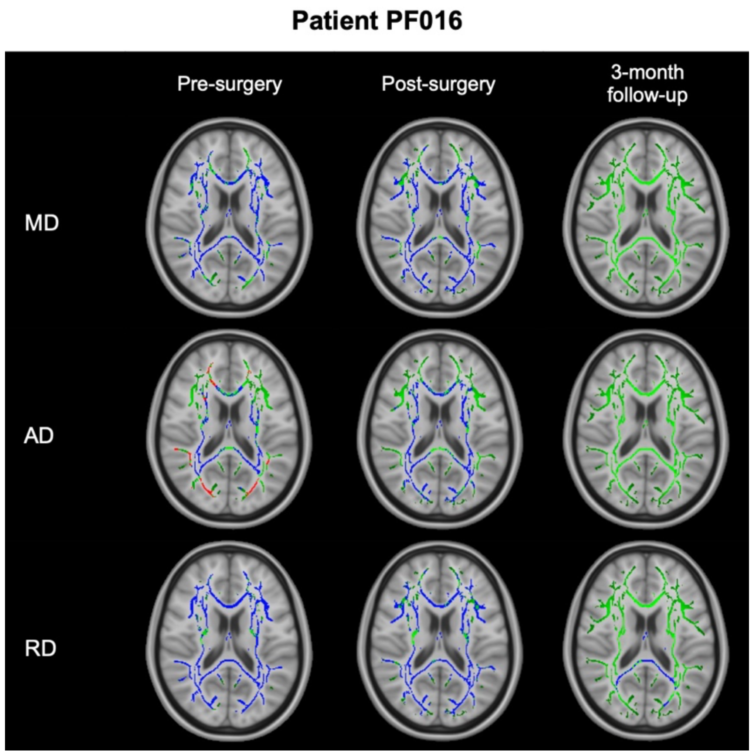


***Figure S6:*** Patient PF016 TBSS results of DTI diffusivity parameters (MD, AD and RD). Green voxels correspond to the white matter skeleton, where voxels were not significantly different between the individual patient and group of controls. Red voxels correspond to areas of the white matter skeleton that were significantly decreased in the patient (p-value < 0.05), and blue voxels were significantly increased in the patient (p-value < 0.05).

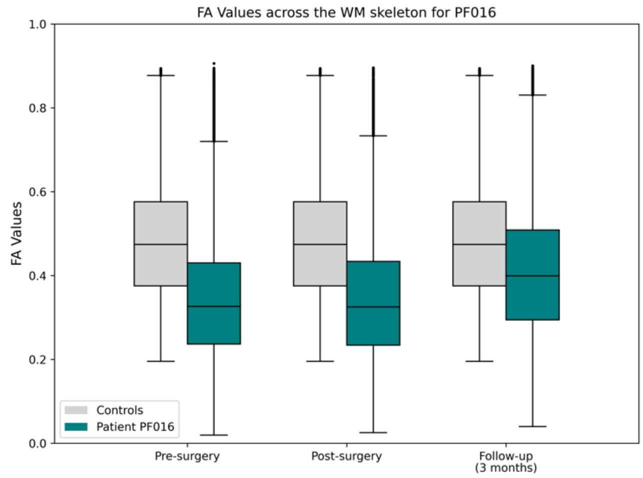


***Figure S7:*** Median FA values across the full white matter skeleton in patient PF016 compared to the average median FA values across the full white matter skeleton in the control group.

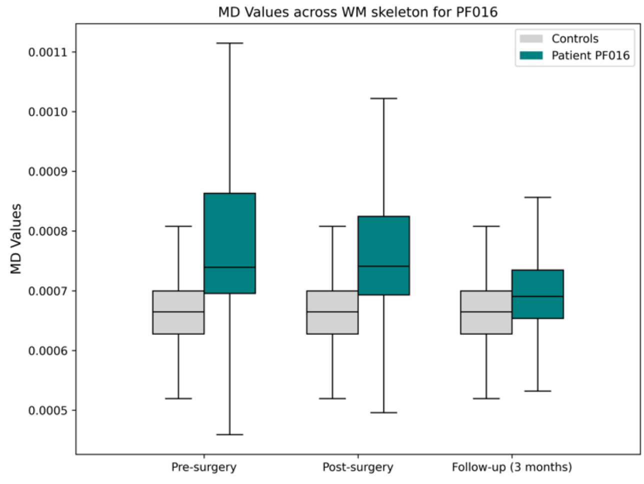


***Figure S8*:** Median MD values across the full white matter skeleton in patient PF016 compared to the average median MD values across the full white matter skeleton in the control group. Outliers have been removed from the box plot for clarity.

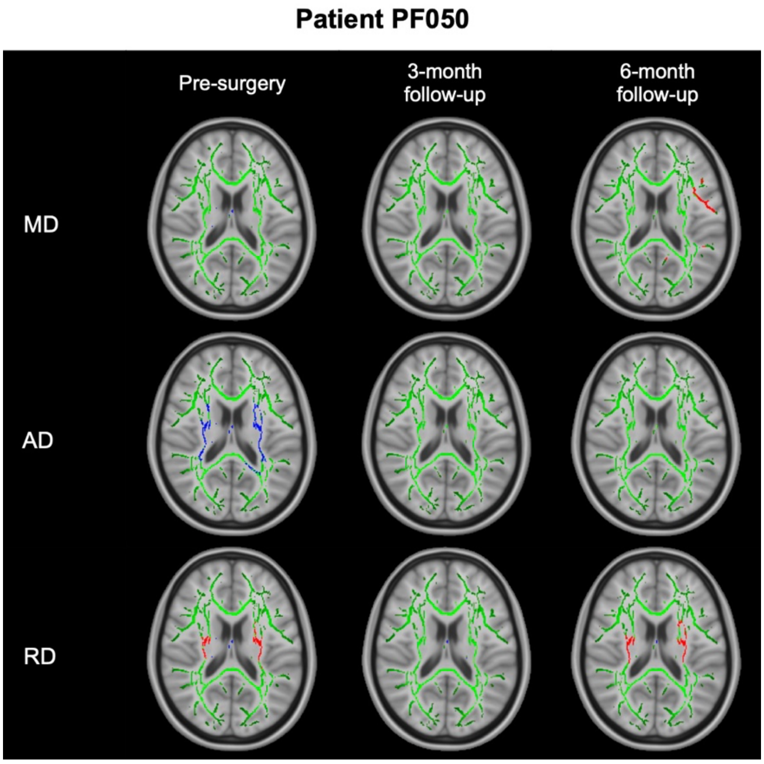


***Figure S9:*** Patient PF050 TBSS results of DTI diffusivity parameters (MD, AD and RD). Green voxels correspond to the white matter skeleton, where voxels were not significantly different between the individual patient and group of controls. Red voxels correspond to areas of the white matter skeleton that were significantly decreased in the patient (p-value < 0.05), and blue voxels were significantly increased in the patient (p-value < 0.05).

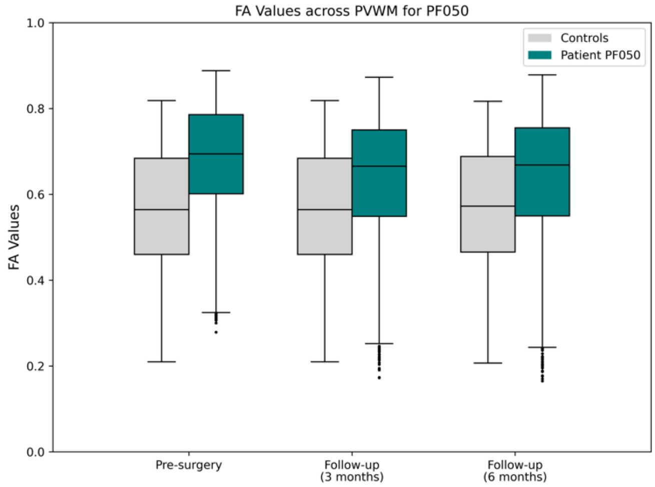


***Figure S10:*** Median FA values from PVWM in patient PF050 compared to the average median FA values from PVWM in the control group.

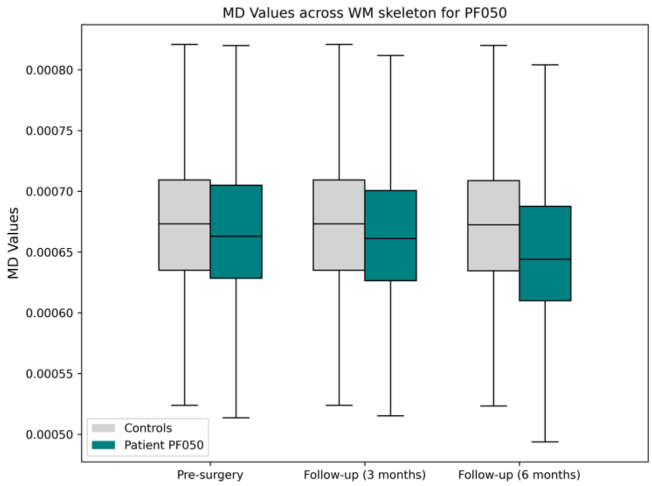


***Figure S11:*** Median MD values across the full white matter skeleton in patient PF050 compared to the average median MD values across the full white matter skeleton in the control group. Outliers have been removed from the box plot for clarity.
